# Factors Influencing the Uptake of Public Health Interventions Delivery by Community Pharmacists: A thematic literature review

**DOI:** 10.1101/2024.01.31.24302091

**Authors:** Audrey Mumbi, Peter Mugo, Edwine Barasa, Gilbert Abiiro, Jacinta Nzinga

## Abstract

**Background:** Community pharmacies are the first point of contact for most people seeking treatment for minor illnesses in Low– and middle-income countries (LMICs). In recent years, the role of community pharmacists has evolved, and they play a significant role in the delivery of public health interventions (PHIs) aimed at health promotion and prevention such as smoking cessation services, weight management services, HIV prevention, and vaccination. This review aims to explore the evidence on the factors that influence community pharmacists to take up the role of delivery of these interventions.

**Methods:** Three electronic databases namely, Embase, Medline, and Scopus were searched for relevant literature from the inception of the database to December 2023. Reference lists of included articles were also searched for relevant articles. A total of 22 articles were included in the review based on our inclusion and exclusion criteria. The data were analyzed and synthesized using a thematic approach to identify the factors that influence the community pharmacist’s decision to take up the role of PHI delivery. Reporting of the findings was done according to the PRISMA checklist.

**Findings:** The search identified 10,927 articles of which 22 were included in the review. The main factors that drive the delivery of PHIs by community pharmacists were identified as; training and continuous education, remuneration and collaboration with other healthcare professionals. Other factors included structural and workflow adjustments and support from the government and regulatory bodies.

**Conclusions:** Evidence from this review indicates that the decision to expand the scope of practice of community pharmacists is influenced by various factors. Incorporating these factors into the design of policies and public health programs is critical for the successful integration of community pharmacists in the delivery of broader public health to meet the rising demand for health care across health systems.

## Introduction

Community pharmacies are the first point of contact with the health system for most people seeking treatment for minor illnesses in Low and Middle-Income Countries(LMICs)(1). They are easily accessible, widely distributed, provide quicker services, open for longer hours, and are relatively cheaper than other private health facilities (1–3). Additionally, they provide a more casual setting for individuals by offering services over the counter for those who do not wish to seek health services from health facilities (4).

Community pharmacists’ role has evolved over the years from the traditional role of distribution and dispensing of prescription and non-prescription medicine to the provision of various Public Health Interventions (PHIs) due to the increased health demands of the population (5). This role has been endorsed by the International Pharmaceutical Federation (FIP) and is recognized in several income high-income countries such as Australia, the United States, United Kingdom it has been integrated into existing healthcare models (6).

Public health entails three main domains; health improvement, health protection, and health service delivery and community pharmacists contribute greatly to all three domains (7). Community pharmacists contribute to improving the health and well-being of the population through provision of a range of services. These include smoking cessation services such as Nicotine Replacement Therapy (NRT) and counseling services (8); provision of interventions aimed at promoting health and well-being through changing of lifestyle habits, healthy weight management services, advice on healthy living, and participation in health promotion campaigns (9, 10). In regards to health protection, community pharmacists offer disease control measures, screening for risk factors for non-communicable diseases such as Cardiovascular Disease (CVD) (11, 12), Sexually Transmitted Infections (STIs) screening (13), Human Immune deficiency Virus (HIV) screening (14), provision of immunization services and communicating information on threats to health to patients and the public in general (7). Health service quality entails the provision of innovative quality pharmacy services to improve health outcomes for instance through medication therapy management services and supporting the safe and effective use of medicine (7). This review mainly focuses on health improvement and health protection domains, as the community pharmacists’ roles are clearly defined.

The delivery of PHIs through community pharmacies not only leads to improved health outcomes, but also reduces socio-economic inequalities by treatment of modifiable risk factors, it also reduces the burden on healthcare systems leading to cost reductions (6, 15). Furthermore, it contributes to the reduction of health inequalities in the population through the promotion of health and well-being messages by encouraging people to adopt healthier lifestyles and take responsibility of their health (7).

Despite the evidence of such benefits, several barriers hinder the optimal engagement of community pharmacists in the delivery of PHIs particularly in LMICs. Understanding the factors that influence the decision of community pharmacists to take up the role is essential for the implementation to be successful. This review therefore aims to explore the factors that influence the community pharmacist’s decision to take up the extended role of PHI delivery.

## Research question

What are the factors that influence community pharmacists’ decision to take up the role of PHI delivery?

## Objectives

The objective of this study was to systematically review available evidence on the factors that influence community pharmacists’ decision to take up the role of delivery of Public Health Interventions.

## Methods

A protocol for our review can be found in the Open Science Framework (16). We conducted this systematic review according to the Preferred Reporting Items for Systematic Reviews and Meta-Analyses (PRISMA) Statement (17) and adhered to the PRISMA checklist (S2 Appendix).

### Study selection

Studies were eligible for inclusion if they: 1) reported on interventions delivered in community pharmacies, which are also referred to as private or retail pharmacies. Community pharmacies refer to generally small to medium-sized businesses providing typical pharmacy services such as filling of prescriptions, over-the counter products and point-of-care (POC) testing or self-testing kits for common diseases. We did not include interventions delivered in hospitals, clinics, or online pharmacies; 2) reported on factors that influence the uptake of the delivery of PHIs. We excluded studies that reported on interventions aimed at antimicrobial resistance, improving treatment and management of diseases, self-medication or management interventions with no screening or diagnosis components; 3) interventions were provided by registered pharmacists and/or pharmacy technicians (in some cases referred to as pharmacy assistants); 4) were published in the English language.

The screening of the identified articles was conducted in two steps:

Step 1: Following the removal of duplicates, the articles were screened based on their title and abstracts using the inclusion and exclusion criteria. Studies that did not meet the criteria were deemed to be irrelevant and excluded.

Step 2: Full articles of the potentially relevant studies were retrieved, and a detailed screening was conducted using the inclusion and exclusion criteria.

The study selection process is illustrated in Fig 1.

**Fig 1.**
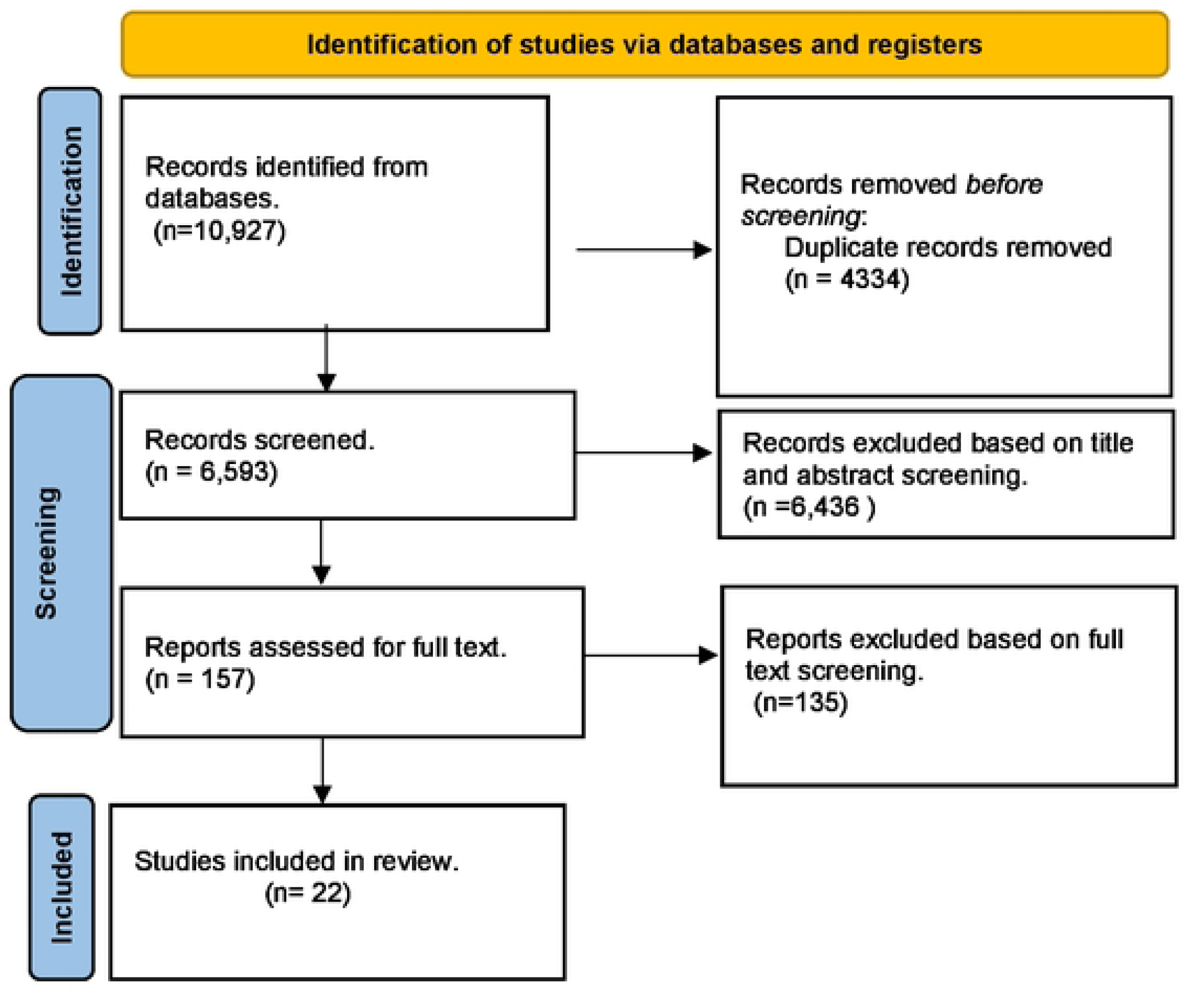

### Search strategy

We searched the literature from July-December 2023 in 3 databases namely: Embase, Scopus, and Medline to identify relevant literature The search terms used were: “Community Pharmacy” “Private pharmacy” OR “Retail pharmacy” AND “preventive health services” OR “public health services” OR “health promotion” OR “screening” OR “testing” OR “Case finding” AND “cardiovascular disease” OR “Diabetes” OR “Blood pressure” OR “drug use” OR “substance use” OR “mental health” OR “sexual health” OR “vaccination” OR “Immunization” OR “Family planning” OR “Contraception”. This search strategy and terms [S1 Appendix] were modified for Embase and Scopus as appropriate. We also searched for relevant literature from reference lists of identified studies. Our search included all published studies from the inception of the databases up to the time of the search. There were no restrictions on the year of publication, study design, or country. The search results from each database were uploaded to End Note 2.0 reference software. The search results from the three databases were collated and duplicates were removed.

### Data Items, Extraction and Analysis

Relevant data from the selected articles were extracted into Ms. Excel by AM with accuracy checks performed on selected articles by JN and any conflict was resolved through discussion of the justification of the inclusion and exclusion criteria. We extracted information on study title, first author, year of study, study country, PHI and factors influencing the uptake of PHI delivery.

The coding process was conducted manually in MS Excel. Data were analyzed using thematic analysis which entailed 4 phases (18). Phase 1: familiarization with the identified articles through reading and re-reading, Phase 2: generation of initial codes that were used to develop the coding framework, Phase 3: coding the contents of the articles onto the coding framework, and Phase 4: generation of themes by identifying patterns and relationships across the identified codes. We used established themes to summarize the findings descriptively and summary tables.

### Quality assessment

The studies were assessed for quality using the Critical Appraisal Skills Programme (CASP) which uses a standardized checklist to assess the adequacy, trustworthiness, and relevance of the evidence reported in the articles (19, 20). We used the Appraisal tool for Cross-Sectional Studies (AXIS tool) to assess for quality of cross sectional studies (21). Tables 2 and 3 outline the results from the quality assessment.

## Results

Our search yielded 10,927 articles from the three databases, of these 4,434 were duplicates, 6,436 were excluded after the screening of the title and abstracts,157 articles were included in the full-text review, and we included 22 studies in the final review (Fig 1).

### Characteristics of the selected studies

Table 1 shows the characteristics of the studies included in the review. Twenty-two studies were selected for the review. Out of the 22, 4 studies were conducted in United Kingdom (UK), 2 in United States of America (USA), 3 in Australia, 4 in Canada, and 1 from Nigeria, Poland, Portugal, Austria, Lebanon, Malaysia, Pakistan, United Arab Emirates (UAE) and Qatar. The main public health interventions identified from the review were; vaccination services (6), healthy weight management (4), Emergency hormonal Contraceptives (EHC) (4), chlamydia screening (2), Cardiovascular screening (CVS) (2), HIV services (2), Health education (1), Pre-exposure prophylaxis screening (PrEP) (1), Diabetes screening (1).

**Table 1:** Characteristics of selected papers.

| **First Author** | **Country** | **Study title** | **Intervention (PHI)** | **Identified factors** |
| --- | --- | --- | --- | --- |
| **L. Ahmaro (22)** | UK | Investigating community pharmacists' perceptions of delivering chlamydia screening to young people: a qualitative study using normalisation process theory to understand professional practice | Chlamydia Screening | Training and continuous education |
| **H. Alzubaidi(23)** | UAE | Pharmacists' experiences and views on providing screening services: An international comparison | CVS risk screening | Remuneration.<br>Structural adjustments |
| **Moore S (24)** | UK | Health promotion in the high street: a study of community pharmacy | Health promotion activities | Training and continuous education<br>Remuneration<br>Organizational adjustments |
| **M. Atif (25)** | Pakistan | A qualitative study to explore the role of pharmacists in healthy weight management in adults in Pakistan: Current scenario and future perspectives | Healthy weight management | Training and continuous education<br>Support from government and Professional bodies |
| **M. Cerbin-Koczorowska (26)** | Poland | Is there a time and place for health education in chain pharmacies? Perspectives of Polish community pharmacists | Health education | Training and continuous education<br>Structural and organizational adjustments<br>Remuneration |
| **B. Chirewa(27)** | United Kingdom | Emergency hormonal contraceptive service provision via community pharmacies in the UK: a systematic review of pharmacists' and young women's views, perspectives and experiences. | Emergency Hormonal Contraceptive | Training and continuous education<br>Collaboration with health care professionals |
| **L. S. Deeks (28)** | Australia | Can pharmacy assistants play a greater role in public health programs in community pharmacies? Lessons from a chlamydia screening study in Canberra, Australia | Chlamydia Screening | Training and continuous education<br>Remuneration |
| **S. Almukdad (29)** | Qatar | Exploring the role of community pharmacists in obesity and weight management in Qatar: A mixed-methods study. | Weight management services | Structural adjustments<br>Training and continuous education<br>Collaboration with health care professionals |
| **W. C. Ang (30)** | Malaysia | Readiness and willingness of Malaysian community pharmacists in providing vaccination services | Vaccination services | Training and continuous education<br>Collaboration with health care professionals<br>Structural and organizational adjustments |
| **L. Chen (31)** | USA | Implementation of hormonal contraceptive furnishing in San Francisco community pharmacies. | Contraceptives | Structural and organizational adjustments |
| **M. Donald (32)** | Canada | Patient, family physician and community pharmacist perspectives on expanded pharmacy scope of practice: a qualitative study | CVD | Training and continuous education<br>Support from professional organization |
| **N. Edwards (33)** | Canada | Pharmacists as immunizers: a survey of community pharmacists' willingness to administer adult immunizations | Immunizations | Training and continuous education |
| **I. Figueira (34)** | Portugal | Point-of-care HIV and hepatitis screening in community pharmacies: a quantitative and qualitative study | Screening for HIV, Hepatitis C & Hepatitis B | Training and continuous education |
| **R. Hopkins (35)** | USA | Support and perceived barriers to implementing pre-exposure prophylaxis screening and dispensing in pharmacies:<br>Examining concordance between pharmacy technicians and pharmacists | Prep screening & dispensing | Training and continuous education<br>Structural adjustment<br>Remuneration |
| **J. E. Isenor (36)** | Canada | Pharmacists' immunization experiences, beliefs, and attitudes in New Brunswick, Canada. | Immunization | Training and continuous education<br>Structural adjustment<br>Remuneration |
| **D. V. Kelly (37)** | Canada | Expanding access to HIV testing through Canadian community pharmacies: findings from the APPROACH study | HIV POCT | Training and continuous education<br>Organizational adjustment<br>Remuneration<br>Collaboration with other health care professional |
| **N. Lindne (38)** | Austria | The willingness of community pharmacists to immunise: A national cross-sectional study | Immunization | Training and continuous education<br>Support from government |
| **K. P. Osemene (39)** | Nigeria | Evaluation of community pharmacists' involvement in public health activities in Nigeria. | Public health interventions ( broad) | Training and continuous education<br>Structural adjustment<br>Collaboration with health care professionals |
| **A. H. Y. Siu (40)** | Australia | Implementation of diabetes screening in community pharmacy - factors influencing successful implementation | Diabetes screening | Training and continuous education<br>Remuneration |
| **I. S. Um (41)** | Australia | Weight management in community pharmacy: What do the experts think? | Weight management | Training and continuous education<br>Collaboration with health care professionals<br>Structural adjustments |
| **A. E. Weidmann (42)** | Scotland | Promoting weight management services in community pharmacy: perspectives of the pharmacy team in Scotland | Weight management | Training and continuous education |
| **D. Youssef (43)** | Lebanon | Exploring determinants of community pharmacist-led influenza vaccination in a Middle Eastern country: a national web-based cross-sectional study. | Immunization | Remuneration<br>Support from government |

### Quality of evidence

All the included studies had clear statements on the aims, methods used, research design and data collection procedures, data analysis and made significant contributions to existing knowledge and discussed the transferability of the findings to other contexts. However, most studies scored poorly on the relationship between the researchers and participants, and for the quantitative studies they scored poorly on measures taken to address non-responders. Despite this, we decided to include all the studies in the review as they adequately met the inclusion criteria and contributed to our review objective. Table 2 and 3 below outline the findings from the quality assessment of the studies using CASP & AXIS tools respectively.

**Table 2:** Assessment of qualitative studies.

| Appraisal criteria | Yes | Somewhat | No/Not clear |
| --- | --- | --- | --- |
| 1. Was there a clear statement of the aims of the research? | 16 |  |  |
| 2. Is a qualitative methodology appropriate? | 16 |  |  |
| 3. Was the research design appropriate to address the aims of the research?<br>Has the researcher justified the research design? | 16 |  |  |
| 4. Is the recruitment strategy appropriate for the study aims?<br>Researcher explained how the inputs/outputs/study participants were selected?<br>Discussion around recruitment, i.e., why some inputs/outputs/participants were not chosen? | 14 | 2 |  |
| 5. Does the data collection approach appropriately answer the research question?<br>• Was the data collection location justified?<br>• If it clear how data were collected?<br>• Were data collection methods clear? | 16 |  |  |
| 6. Has the relationship between the researcher and participants adequately been considered?<br>• Researcher reflexivity and potential partiality during the formulation of research questions or data collection? | 2 |  | 14 |
| 7. Did the researcher consider ethical issues before conducting the study?<br>• Are issues on informed consent and confidentiality adequately addressed?<br>• Did the researchers seek ethical approval? | 14 |  | 2 |
| 8. Was there adequate rigor during data analysis?<br>• An explicit explanation of how the analysis was conducted.<br>• A clear statement of how themes/categories were developed.<br>• Are there proper considerations to inconsistent findings? | 14 | 2 |  |
| 9. Are the findings reported clearly?<br>• Explicit findings<br>• An adequate discussion of evidence for and against the researcher arguments<br>• The credibility of finds (triangulation, respondent validation, more than 1 analysis) findings are discussed in relation to the original research question | 16 |  |  |
| 10. How valuable is the research?<br>• The researcher explains how the study contributes new knowledge or adds to existing knowledge.<br>• Have the research identified new areas for future research.<br>• Are there clear explanations about how the findings can be applied to other settings. | 16 |  |  |

**Table 3:** Assessment of Cross sectional studies.

| **AXIS-Cross sectional studies checklist** | **Yes** | **Somewhat** | **No/Not clear** |
| --- | --- | --- | --- |
| Were the aims /objectives of the study clear? | 6 |  |  |
| Was the study design appropriate for the stated aim? | 6 |  |  |
| Was the sample size justified? | 6 |  |  |
| Was the target/reference population clearly defined? (Is it clear who the research was about?) | 6 |  |  |
| Was the sample frame taken from an appropriate population base so that it closely represented the target/reference population under investigation? | 5 | 1 |  |
| Was the selection process likely to select subjects/participants that were representative of the target/reference population under investigation? | 6 |  |  |
| Were measures undertaken to address and categorize non-responders? |  |  | 6 |
| Were the risk factor and outcome variables measured appropriate to the aims of the study? | 6 |  |  |
| Were the risk factor and outcome variables measured correctly using instruments/measurements that had been trialed, piloted or published previously? | 4 | 2 |  |
| Is it clear what was used to determine statistical significance and/or precision estimates? (e.g., p-values, confidence intervals) | 6 |  |  |
| Were the methods (including statistical methods) sufficiently described to enable them to be repeated? | 6 |  |  |
| Were the basic data adequately described? | 6 |  |  |
| Does the response rate raise concerns about non-response bias? |  |  | 6 |
| Were the results internally consistent? | 6 |  |  |
| Were the results presented for all the analyses described in the methods? | 6 |  |  |
| Were the author's discussions and conclusions justified by the results? | 6 |  |  |
| Were the limitations of the study discussed? | 4 |  | 2 |
| Were there any funding sources or conflicts of interest that may affect the authors' interpretation of the results? |  |  | 6 |
| Was ethical approval or consent of participants attained? | 6 |  |  |

### Synthesis of results

We identified 6 major themes from this review: training and education (19/22), structural and organizational adjustments (11/22), remuneration (9/22), collaboration with other health care professionals (6/22), support from government and professional bodies (4/22). These findings have been summarized in table 4 below.

**Table 4:** Factors influencing PHI delivery.

| **Factors** | **Country** | **Studies** |
| --- | --- | --- |
| **Training and continuous education** | UK<br>Pakistan<br>Poland<br>Australia<br>Qatar<br>Malaysia<br>Canada<br>Portugal<br>USA<br>Nigeria | L. Ahmaro et al (22)<br>Moore S et al (24)<br>M. Atif et al (25)<br>M.Cerbin et al (26)<br>B. Chirewa et al (27)<br>L.S.Deeks et al (28)<br>S. Almukdad et al (29)<br>W.C.Ang et al (30)<br>M. Donald et al (32)<br>N.Edwards et al (33)<br>I. Figueira et al (34)<br>R. Hopkins et al (35)<br>J.E. Isenor et al (36)<br>D.V.Kelly et al (37)<br>N.Lindne et al (38)<br>K.P.Osemene et al (39)<br>A.H.Y.Siu et al (40)<br>I.S.Um et al (41) |
| **Remuneration** | UAE<br>UK<br>Poland<br>Australia<br>USA<br>Canada<br>Lebanon | H. Alzubaidi et al (23)<br>Moore S et al (24)<br>M.Cerbin et al (26)<br>L.S.Deeks et al (28)<br>R. Hopkins et al (35)<br>J.E.Isenor et al (36)<br>D.V.Kelly et al (37)<br>A.H.Y.Siu et al (40)<br>D. Youssef et al (44) |
| **Structural and organizational improvements** | UAE<br>UK<br>Poland<br>Qatar<br>Malaysia<br>USA<br>Canada<br>Nigeria<br>Australia | H. Alzubaidi et al (23)<br>Moore S et al (24)<br>M.Cerbin et al (26)<br>S. Almukdad et al (29)<br>W.C.Ang et al (30)<br>L. Chen et al (31)<br>R. Hopkins et al (35)<br>J.E.Isenor et al (36)<br>D.V.Kelly et al (37)<br>K.P.Osemene et al (39)<br>I.S.Um et al (41) |
| **Collaboration with health care professionals** | UK<br>Qatar<br>Canada<br>Nigeria<br>Australia | B. Chirewa et al (27)<br>S. Almukdad et al (29)<br>W.C.Ang et al (30)<br>D.V.Kelly et al (37)<br>K.P.Osemene et al (39)<br>I.S.Um et al (41) |
| **Government support** | Pakistan<br>Canada | M. Atif et al (25)<br>M. Donald et al (32) |
|  | Austria<br>Lebanon | N.Lindne et al (38)<br>D. Youssef et al (44) |

### Training and Continuous Education

Pharmacists are qualified to provide health care services, however, the reviewed literature suggests that they feel incompetent in taking up this role and would benefit from additional training on the provision of specific PHIs (22) (24–30, 32–42). Pharmacists in the UK received training on how to deliver chlamydia testing by attending a sexual health learning training session and through the completion of an online learning module. (22). Elsewhere, Moore S et al found that 70% (21/30) of pharmacists acquired health promotion training in their undergraduate degree and informally through reading articles in professional journals rather than attending a formal training program (24). In Canada, vaccination training was provided to pharmacists through online learning modules and live day training (36). Almukdad et al, exploring the attitudes of pharmacists in the role of offering weight management services found that pharmacists would benefit from regular and structural training to promote their expertise (29). Pharmacists in Malaysia reported they would prefer to have at least 1 or 2 days of short training workshops (30).

Pharmacists in Pakistan and Poland expressed a gap in formal training in the undergraduate curriculum and they recommended the inclusion of more practical training sessions in the curriculum (25, 26). They also expressed concern about the high fees of the courses, and they suggested the need for funding opportunities for free courses. Community pharmacists in Australia and Malaysia reported willingness to provide vaccination services after having received additional short training that covers aspects of needle size gauge and landmarking (36), vaccine storage, technique, and handling of emergency cases (30), with a 2 year renewal interval (38). Training was suggested for both pharmacists and their assistants in 4 studies to facilitate task shifting of responsibilities (24) (28) (39) (36).

### Remuneration

The provision of PHIs was viewed as an additional workload on top of the dispensing and pharmaceutical-based services and remuneration is a pre-requisite for further activity (23) (24) (26) (28) (29) (30) (35) (36). Pharmacists in Poland expressed they would like to be compensated through an increase in salary to motivate them to take up the role of a health educator (26). Elsewhere, pharmacists in the UK reported they would prefer to be compensated through a fee-for-service model by the Kingston & Richmond Family Health Service Authority (FHSA) (24). Pharmacy assistants in Australia viewed remuneration as a tangible recognition for the provision of Chlamydia screening and participation in the training program, in addition to receiving a certificate (28). None of the other studies reported on the modes of remuneration but there was consensus that remuneration was a means of motivating pharmacists. For instance, pharmacists in the UAE reported that remuneration would motivate them to screen more clients for STIs (23). There was also an agreement among pharmacists in Australia and the UAE that screening services should be offered at minimal or no costs, funded through government subsidies (23).

### Structural and workflow adjustments

Structural adjustments such as the establishment of a dedicated space for private consultation and as an enabler for the adoption of this role. This was indicated in 8 studies; (24) (25) (26) (29) (30) (31) (34) (39). For example, community pharmacists in Nigeria reported that having a designated space for offering patient counseling would encourage them to participate in more public health services (39), similar to findings by Almukdad et al where pharmacists proposed having a private area for counseling as an internal strategy to improve the provision of weight management services (WMS) (29). The need for a designated space with proper equipment to measure obesity-related parameters for screening blood pressure, weight measurement and cholesterol measurement was reported in Pakistan (25). For provision of vaccination services pharmacists expressed the need for a specified area for provision of the vaccine services and storage of vaccines under the right temperatures (30) (36). However, none of the studies detailed how this could be achieved.

Elsewhere organizational adjustments such as having a time dedicated time for delivery of PHIs to avoid disruption of the normal dispensing activities (24) (26) (30) (31) (35) was reported as an enabler for uptake of PHI delivery. Providing training to pharmacy assistants on how to handle pharmacy-based activities was reported as a strategy to free up time for pharmacists to be involved in the delivery of PHIs (39) (26), as well as scheduling appointments for the PHIs (31).

### Collaboration with other healthcare professionals

Pharmacists expressed that the provision of additional services (i.e. delivery of PHIs) would require a multidisciplinary approach (24) (25) (32) (41) (29), especially for cases that required referral and consultation with other health care professionals such as obesity cases (41). Pharmacists reported they would prefer having multi-disciplinary training courses as a way of forming links between different professional groups (24). Pharmacists in Canada expressed low awareness among general physicians on pharmacists’ capabilities in providing PHIS and suggested having a shared electronic medical record with physician to facilitate an integrated model of care (32). Elsewhere in Qatar, pharmacists viewed offering Weight Management Services as complex and they highlighted the need for collaboration with dieticians and physicians for referral purposes (29). However, the findings from these studies were mainly aspirational, and none reported on an existing collaboration model.

### Support from the government and professional bodies

Support from governing bodies was identified as crucial in the implementation of public health programs in four studies (25, 29, 30) (32). For example, in Pakistan pharmacists reported there was low awareness of the role pharmacists play in public health and the government could play a role in promoting awareness to the public which in turn would enhance trust from the public (25). These findings were similar in Canada where pharmacists expressed that professional associations could play a role in creating awareness of this role (32). Pharmacists in Qatar expressed the need for the Ministry of Health to establish guidelines for pharmacists to facilitate the adoption of Weight Management Services (WMS) (29). Elsewhere in Malaysia, pharmacists expressed that the government could support them in this role by offering free training and resources to facilitate the vaccination role and professional bodies by advocating for the roles of CPs to be included in vaccination programs (30).

## Discussion

To the best of our knowledge, this is the first review highlighting the factors that influence community pharmacists to expand their scope of practice and deliver PHIs from a global perspective. The findings from this review reaffirm that pharmacists are willing to expand their practice beyond dispensing and take up the role of PHI delivery.

The core finding of this review was that offering additional training to community pharmacists on the delivery of specific PHIs is a requirement to boost the uptake of this role. Training could augment pharmacists’ knowledge and skills as well as empower them to be more competent and confident in the delivery of PHI. The benefits of training pharmacists on PHI delivery include elevating their confidence and competence in service delivery and thus improved health outcomes (45–47). There was no reported standardized model in place for training pharmacists on PHI delivery from the studies reviewed. Nevertheless, various training models have been adopted elsewhere although these differ by context and the PHI under consideration.

These training models include peer learning, which has the potential to influence the practitioner’s behavior (48), learning at work (49) (50), and formal certification to become specialists (51). These trainings are delivered through different formats such as face-to-face learning ( onsite/off-site), and online learning (webinars-learning modules and activities). Face-to-face training has been reported as the preferred mode of delivery by pharmacists in Ethiopia (49), UAE (52), and USA (53), as it offers an opportunity for quick feedback from the instructor, provides an opportunity for peer networking, which could enhance the collaboration with health care professionals in other fields. Online training on the other hand has been reported as a preferred mode of delivery by pharmacists in Australia (54), as it offers the convenience of pharmacists completing modules at their own pace and schedule. Blending diverse training methods and modes by pharmacists’ preferences is key to ensuring that pharmacists are well-equipped to take up the role of delivery of PHI.

Governments can offer support to pharmacists by offering free training sessions as well as providing access to courses, workshops, and conferences online. Additionally, they can offer resources to facilitate the adoption of PHI delivery through, for example, the provision of materials to promote awareness and offering equipment at a subsidized price that is affordable to pharmacists (55). The vital role that community pharmacies play in improving health indicators is recognized globally, however, their inclusion in the design of policies, countries’ health strategies regulation and monitoring is minimal (56). The government can therefore play a role by establishing clear guidelines, policies, and regulatory frameworks to guide the integration of this role into broader health systems. Furthermore, the government and professional bodies could promote national campaigns to create awareness of the crucial role that pharmacists play in public health.

Whilst integrating community pharmacist PHI delivery role into the broader health system, it’s important that they generally operate independent of other healthcare providers in a retail environment (57). Therefore, training sessions could create an avenue to foster interprofessional collaborations between pharmacists and other healthcare professionals. Interprofessional collaboration between physicians and pharmacists has resulted in improved patient outcomes and a reduction of health system inefficiencies and costs (58) (59). This has led to the establishment of a range of initiatives such as Collaborative Practice Agreements (CPAs) in the USA to allow collaborative medication therapy management (60), the establishment of pharmacotherapy groups between General Physicians (GPs) and pharmacists in the Netherlands (61), and home medicine reviews in Australia (62). Interprofessional collaboration has been described as an evolving process that progresses through a series of stages described by various collaboration models. For instance, a GP-pharmacist model by McDonough and Doucette (63) describes it as a progression in 4 main stages: stage 0-professional awareness, stage 1-professional recognition, stage 2-exploration and trial stage, stage 3-professional relationship expansion, stage 4-a commitment to the collaborative working relationship and is influenced by different factors such as proximity, time, clinical knowledge, communication, mutual interests and professional equality. Other models have been adopted for collaboration between pharmacists and GPs and are similar in that collaboration progresses from brief interactions to a clearly defined relationship where the roles of both cadres are well defined (64, 65). Role clarity has been shown to influence the adoption of role expectations and task performance (66, 67).

Community pharmacies are private retail businesses operating within a competitive market and aim to maximize profits to survive in the market, it is therefore not surprising that remuneration influences the uptake of the additional role. Delivery of PHI is viewed as an additional role and pharmacists have few incentives to deliver the expanded services if the compensation is inadequate. Although the remuneration of community pharmacists has mainly been based on their retailing and dispensing functions, a few countries have introduced payment mechanism reforms as a means of encouraging the adoption of this role (68). For instance, the fee-for-service model has been adopted to encourage pharmacists to provide smoking cessation services (69), influenza vaccination (70), and diabetes-related education, training and monitoring in the community settings (71) and was more preferred by pharmacists as it was easy to implement and integrate into the existing business model (72). Pay-for-performance model has been used in a UK program, where pharmacies were renumerated based on the number of people who successfully quit smoking (73). However, there is a gap in knowledge on the preferred payment model in various contexts. Understanding the payment model preferences of community pharmacists is a crucial knowledge gap as it has major implications on the implementation, adoption, and potential impact of pharmacists’ payment model.

Finally, structural and workflow adjustments such as having a designated space and having a dedicated time play a role in community pharmacists taking up the role of PHI. The importance of a private room has been stressed in several studies as a way of building trust and maintaining confidentiality for patients who want to discuss sensitive medical issues such as requests for EHC, screening for STIs, Prep, and HIV screening (74) (75). Evidence suggests that community pharmacists have a preference for having a private consultation room to provide services for diabetes management to preserve patients’ privacy and confidentiality (76). This can be achieved through the establishment by development of policies and standards for the physical space of pharmacies such as having a designated space for conducting PHIs. For instance, having a private space has been incorporated as requirement in Western Australia section 7 of Pharmacy Regulations 2010, which specifies that, “ *The premises are to have an area in which a consultation conducted by a pharmacist is not reasonably likely to be overheard by a person not a party to the consultation”* (77). These guidelines are backed up by the professional body code of ethics as a means to ensure that the client’s right to privacy and confidentiality is maintained (78). Workflow adjustments such as having time dedicated to PHIs would allow pharmacists to plan efficiently and allocate sufficient time to deliver high-quality services. However, evidence shows that community pharmacists have a general preference for being easily accessible to patients by taking walk-in clients (76).

### Limitations

One limitation of this review was that most of the findings from were mainly aspirational and therefore minimal data on various mechanisms that have been applied to facilitate the role of PHI delivery by community pharmacists, however, this will be addressed by a broader study. Second, the findings from this review were mainly in studies conducted in high-income countries and therefore the findings may not be contextually replicable in low-income settings as factors vary across different contexts. To overcome these limitations, further empirical work in LMIC settings is required to determine the key drivers, policy, and practical considerations for delivery of PHIs. Despite these limitations, this manuscript provides crucial information that has great potential to inform the design of public health policies targeting community pharmacists.

### Study implications

This review highlights the different factors that play a key role in influencing community pharmacists’ decision to take up the role of PHI delivery. However further research is needed to generate evidence on how these factors interact to influence implementation practices and sustainability; for two reasons. First, it will ensure that policies are designed in a manner that incentivizes community pharmacists to take up this role. Second, it will facilitate the establishment of guidelines to standardize community pharmacy practice and integration of this role into broader health systems which will in turn enhance the contribution of community pharmacists to public health.

## Conclusion

This review sheds light on the various factors that influence the decision of community pharmacists to expand the scope of practice and take up the role of delivery of public health interventions. Incorporating these factors into the design of policies and public health programs is crucial for the successful integration of community pharmacists into broader public health initiatives. However, these findings do not indicate the relative importance that is placed on each of the factors by community pharmacists. The findings from this review will inform the design of a discrete choice experiment to elicit context-specific preferences of community pharmacists for the identified factors, which will in turn contribute to the design of policies that will enhance the contribution of community pharmacists to public health.

## Authors contributions

This study was conceptualised and designed by AM, PM, EB and JN. AM screened articles, extracted and analysed the data; with accuracy checks performed by JN. All authors were involved in the review and approval of the final submitted version of the manuscript for this work.

## Data Availability

All relevant data are within the manuscript and its supporting information files.

